# SARS-CoV-2 detection by extraction-free qRT-PCR for massive and rapid COVID-19 diagnosis during a pandemic

**DOI:** 10.1101/2020.09.10.20191189

**Authors:** Diana Avetyan, Andranik Chavushyan, Hovsep Ghazaryan, Ani Melkonyan, Ani Stepanyan, Roksana Zakharyan, Varduhi Hayrapetyan, Sofi Atshemyan, Gevorg Martirosyan, Gayane Melik-Andreasyan, Shushan Sargsyan, Armine Ghazazyan, Naira Aleksanyan, Xiushan Yin, Arsen Arakelyan

## Abstract

COVID-19 pandemic severely impacted the healthcare and economy on a global scale. It is widely recognized that mass testing is an efficient way to contain the infection spread as well as the development of informed policies for disease management. However, the current COVID-19 worldwide infection rates increased demand in the rapid and reliable screening of SARS-CoV-2 infection.

We compared the performance of qRT-PCR in direct heat-inactivated, heat-inactivated/pelleted samples against RNA in a group of 74 subjects (44 positive and 30 negative). In addition, we compared the sensitivity of heat-inactivated/pelleted in another group of 196 COVID-19 positive samples.

Our study suggests that swab sample heat-inactivation and pelleting show higher accuracy for SARS-CoV-2 detection PCR assay compared to heat-inactivation only (89% vs 83% of the detection in RNA). The accuracy of detection using direct samples varied depending on the sample transport and storage media as well as the concentration of viral particles.

Our study suggests that purified RNA provides more accurate results, however, direct qRT-PCR may help to significantly increase testing capacity. Switching to the direct sample testing is justified if the number of tests is doubled at least.

## Introduction

Coronavirus disease (COVID-19) is a respiratory tract infection caused by severe acute respiratory syndrome coronavirus 2 (SARS-CoV-2) [1]. It was first identified in Wuhan, Hubei Province, China, in December 2019, and the genetic sequence of the novel coronavirus was already shared in January 2020 [2], [3]. The current COVID-19 worldwide infection rates and a high proportion of asymptomatic cases forced an unexpected burden on the health care systems worldwide and led to massive, rapid, affordable, and efficient diagnostic tests for daily screening.

The current standard method for the detection of RNA viruses in clinical diagnostics is based on the extraction of RNA from a nasopharyngeal swab in viral transport media (VTM) followed by reverse transcription quantitative polymerase chain reaction (qRT-PCR) [4]. Additionally, the existing protocol was optimized to one-step qRT-PCR, which removes the need for a separate reverse-transcription (RT) step resulting in complementary DNA (cDNA) synthesis from the viral RNA template [5]. However, in many sites, automatic isolation is not available, while manual isolation takes 2-4 hours and requires significant work that can result in additional experimental errors as well as limits the possibilities to extend testing capacity. On the other hand, there is also a risk of reagent shortage in major kit suppliers. Therefore, the ability to omit the RNA purification from diagnostic protocols would be not only affordable, operative, and efficient for COVID-19 screening, but also keeps the use of commercial kits to the minimum. The options of detection of viral RNA by qRT-PCR directly from VTM, UTM (Universal Transport Medium) media have been already explored [4–8]. The results are promising, however, it has been shown that Ct values in those samples are shifted compared to the Ct values from purified RNA samples, which could cause a decrease of sensitivity. And as a consequence, the “false-negative” results increase. Here we optimized two different protocols for qRT-PCR with direct samples and systematically compared them with the current detection assay with RNA extraction.

## Materials and methods

### Samples

We collected paired nasopharyngeal swabs and RNA samples from 270 patients. The first group of 76 samples was collected randomly, without knowing the sample status. The second group of 196 samples was collected from SARS-CoV-2 positive patients only. The RNA and nasopharyngeal swab samples were obtained from the National Center for Disease Control and Prevention of MH RA, National Centre For Aids Prevention of MH RA, and Davidyants Laboratories. Swab samples were stored in three different transport media types: Sample Storage Reagent (Sansure Biotech, China) (n = 45) media, PBS (n = 91) and STORE-F UTM (DNA technologies, Russia) (n = 134). No additional clinical or demographic information was collected. The study was approved by the Institutional Review Board of the Institute of Molecular Biology NAS RA (IRB#: 00004079).

### RNA extraction

RNA extraction was performed using both automated and manual extraction protocols. Automated extraction (45 samples) was performed using Maxwell RSC Viral Total Nucleic Acid Purification Kit (Promega Corporation Inc, US). Manual extraction was performed using a triazole based PREP-NA kit (87 samples) (DNA Technologies Ltd, Russian Federation) and magnetic bead-based ZipPrime nucleic acid isolation kit (64 samples) (ZipPrime Ltd, Turkey) according to the manufacturer’s manuals.

### Direct sample with heat-inactivation

To skip RNA extraction, 20 μl of swab samples in transport media were heat-inactivated at 95 °C for 5 min before loading into the PCR.

### Direct sample with inactivation and pelleting

To skip RNA extraction, 100-300 μl of swab samples in transport media were heat-inactivated at 95 °C for 5 min and centrifuged for 10 min at 12000 g. The supernatant was aspirated to leave 20-30 μl liquid in a tube. Pellet was mixed by pipetting or vortexing and used in PCR.

### qRT-PCR

We performed real-time qRT-PCR targeting the N gene and ORF1ab gene in the conserved region of the new coronavirus genome [2]. The final reaction volume consisted of 10 μl PCR reaction mix (Vazyme Ltd, China), 1.5 μl primer/probe mix (ORF-F: 5′-CCCTGTGGGTTTTACACTTAA-3′ (2 µM final concentration); ORF-R: 5′-ACGATTGTGCATCAGCTGA-3′ (2 µM final concentration); ORF-P: 5′-FAMCCGTCTGCGGTATGTGGAAAGGTTATGG-BHQ1-3′ (0.5 µM final concentration); N-F: 5′-GGGAGCCTTGAATACACCAAAA-3′ (2 µM final concentration); N-R: 5′-TGTAGCACGATTGCAGCATTG-3′(2 µM final concentration); N-P: 5′-HEX-AT CACATTGGCACCCGCAATCCTG-BHQ2-3′ (0.5 µM final concentration)), 5 μl template (sample, negative or positive control) in 20 μl of final reaction volume. Thermal cycling was performed as follows: 55 °C for 5 min (reverse transcription), followed by 94 °C for 2 min and 45 cycles of 94 °C for 5 s, 55 °C for 10 s. A final cooling step at 25 °C for 20 s was also included. Thermal cycling took 1 h and 10 min in total. The protocol was described in detail elsewhere [6]. Samples were considered positive when a signal was detected at Ct < 40 for any gene.

### Data analysis

Accuracy, sensitivity, and specificity for PCR assays were calculated using *caret* and *epiR* packages available in the R software environment for statistical computing. The Wilcoxon matched rank test was used to compare differences in Ct values in matching samples.

## Results and discussion

For this part of the study, we performed qRT-PCR assay for SARS-CoV-2 using matched RNA, heat-inactivated (H), and heat-inactivated and pelleted (HC) samples from 74 patients. Nasopharyngeal swabs were stored in PBS (n = 47) and STORE-F UTM (n = 27) for 12-72 hours at +4 °C. The RNA extraction method was compared to the direct addition of samples to the qRT-PCR with and without centrifugation. The results suggest that the centrifugation step considerably improved accuracy and sensitivity compared with the heat-inactivation alone (Table 1).

**Table 1.** COVID-19 detection qRT-PCR results using direct sample and purified nucleic acid.

|  |  | RNA |  | Performance evaluation |
| --- | --- | --- | --- | --- |
|  |  | Positive | Negative |  |
| HC | Positive | 39 | 3 | Accuracy: 89% (95% CI: 80 – 95%)<br>Sensitivity: 89% (95% CI: 75 – 96%)<br>Specificity: 90% (95% CI: 73 – 98%) |
|  | Negative | 5 | 27 |  |
| H | Positive | 35 | 3 | Accuracy: 83% (95% CI: 74 – 91%)<br>Sensitivity: 80% (95% CI: 65 – 90%)<br>Specificity: 90% (95% CI: 73 – 98%) |
|  | Negative | 9 | 27 |  |

The low sensitivity values in both HC and H could be partially attributed to the issues of detection in samples with low concentration of viral particles close to the detection limit. Interestingly, in these samples, Ct values were mostly detected only by one channel. In fact, in samples with RNA Ct values < 34, the sensitivity for heat-inactivation was 93% (28/30) and for heat-inactivation and pelleting was 97% (29/30). Only one sample with Ct value < 20 was missed using HC samples compared to purified RNA.

On the other side, we report three positive HC samples that hadn’t been detected with the extracted nucleic acid. These samples were positive with Ct values around 22 (not detected in RNA), 32 (Ct > 40 in RNA), and 35 (Ct > 40 in RNA). These samples were also positive in heat-inactivation treatment alone, however, with higher Ct values compared to HC. We suggest that this discrepancy could be a result of a failure during RNA isolation steps using manual extraction protocol.

We also compared the differences between Ct values (ΔCt) obtained in purified RNA with Ct values obtained in heat-inactivated and heat-inactivated and pelleted samples (Table 2). In agreement with the previous studies, we also observed a considerable shift of Ct values towards higher values in direct samples. Meanwhile, the median ΔCt was lower by 1.55 and 2.29 cycles (Wilcoxon signed-rank test p = 0.0018 and < 0.0001 for FAM and HEX channels, accordingly) in HC samples compared to H samples (Figure 1A). We also observed cases where Ct values for HC and H samples were smaller compared to the detection in RNA samples (Figure 1B), which indicates that the omitting of RNA processing steps can reduce degradation. Thus, our results show that additional centrifugation and pelleting steps can improve the sensitivity of the detection of viral nucleic acids without much affecting the workflow of the direct PCR.

**Table 2.** Change in Ct value between qRT-PCR with the direct and purified nucleic acid.

| SARS-CoV-2 RNA conc. | Median ΔCt | | Range ΔCt | |
| --- | --- | --- | --- | --- |
| Heat | FAM | HEX | FAM | HEX |
| High (Ct <25) | 6.51 | 8.35 | 3.81 – 14.84 | 6.22 – 15.16 |
| Intermediate (Ct 25-34) | 6.2 | 7.16 | -6.44 – 11.95 | -4.29 – 12.38 |
| Low (Ct >34) | 3.48 | -3.36 | 2.39 – 8.31 | -1.11 – 7.82 |
| Heat + pelleting |  |  |  |  |
| High (Ct <25) | 5.54 | 6.83 | 2.31 – 13.61 | 3.7 – 13.36 |
| Intermediate (Ct 25-34) | 4.64 | 5.25 | -8.27 – 12.41 | -6.41 – 9.83 |
| Low (Ct >34) | -3.5 | -5.49 | -9.9 – -1.58 | -7.96 – 3.26 |

**Figure 1.**
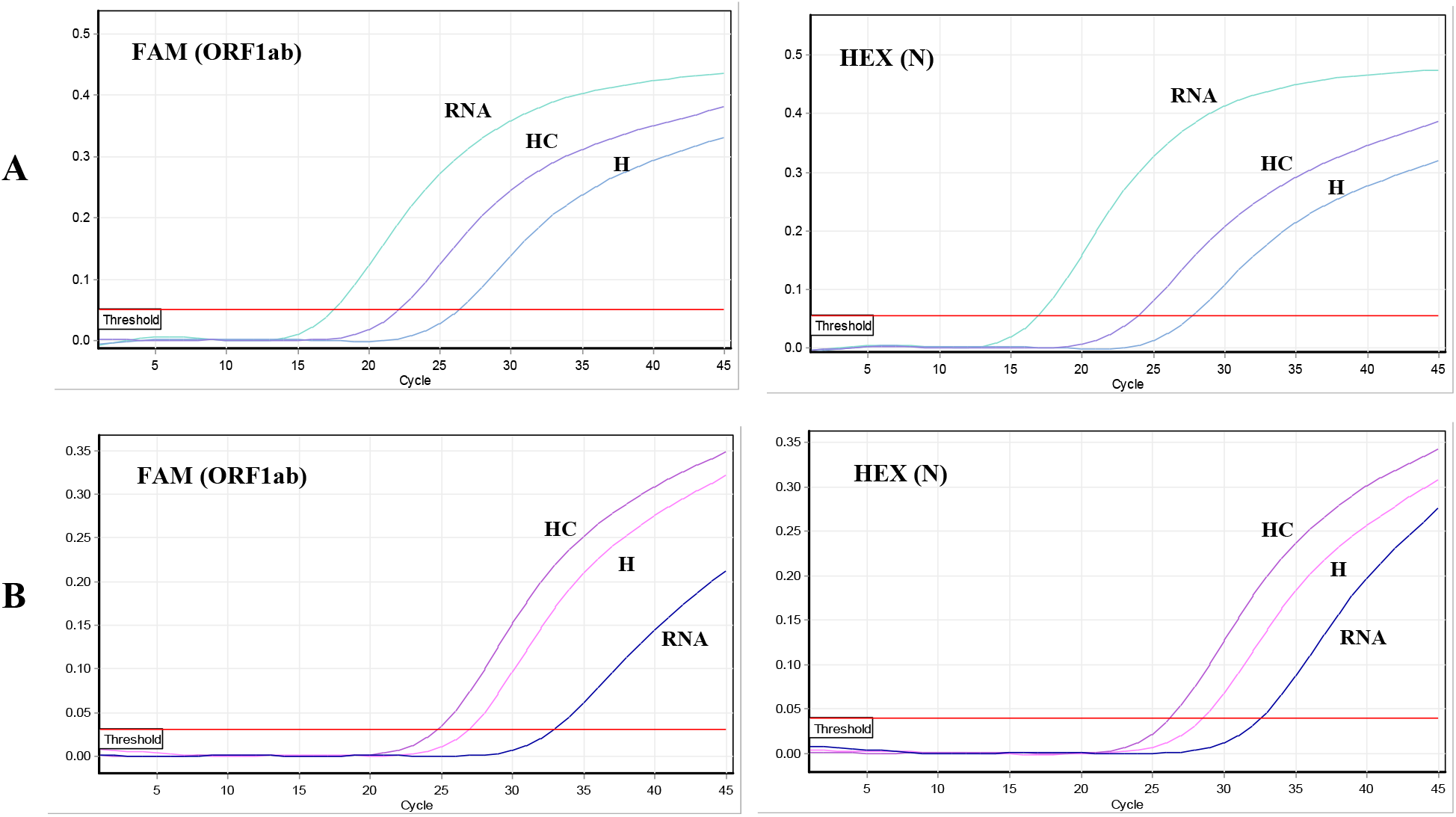
qRT-PCR amplification plots for the SARS-CoV-2 ORF1ab (FAM) and N (HEX) genes for a nasopharyngeal swab sample subjected to the standard RNA extraction, heat inactivation (H), and heat inactivation and pelleting (HC). Amplification plots show normalized value (ΔRn, linear scale) as a function of the qPCR cycle. The horizontal red line denotes the cycle threshold.

Next, we compared the performance of SARS-CoV-2 PCR detection in heat-inactivated (H) and pelleted (HC) samples with the detection in RNA in the larger group of COVID-19 positive samples (Table 3). From 196 samples the HC was able to detect virus 171 samples with the overall sensitivity of 88% (CI95%′ 83% – 92%).

**Table 3.** Detection rates in RNA and HC samples.

|  | **ORF1ab and N** | **ORF1ab only** | **N only** | **Total** |
| --- | --- | --- | --- | --- |
| **RNA** | 174 | 9 | 13 | 196 |
| **HC** | 145 | 14 | 12 | 171 |

Further analyses showed that detection sensitivity varies depending on both transportation media and concentration of viral particles. In general, the sensitivity of detection in HC samples increases along with the increase of viral particle concentrations (Table 4). Moreover, the sensitivity of detection in HC samples differs depending on the transport media in which a sample was stored. The best performance was obtained with Sample Storage Reagent (Sansure Biotech, China) media, followed by PBS and STORE-F UTM (DNA technologies, Russia). However, it should be also mentioned that sample storage time and conditions were not uniform before PCR assay, and we were not able to obtain exact information for each sample. These factors could also affect the performance of the direct assay.

**Table 4.** The sensitivity of direct qRT-PCR with heat-inactivation and pelleting according to the transportation media used and concentration of viral particles.

|  |  | ORF1ab |  |  |  |  | N |  |  |  |  |
| --- | --- | --- | --- | --- | --- | --- | --- | --- | --- | --- | --- |
| | Ct | RNA | HC | Sensitivity | Mean ΔCt | SD ΔCt | RNA | HC | Sensitivity | Mean ΔCt | SD ΔCt |
| **STO R-F** | < 25 | 46 | 41 | 89% | 6.31 | 4.1 | 46 | 38 | 83% | 7.35 | 4.49 |
|  | 25-34 | 20 | 16 | 80% | -0.23 | 8.08 | 20 | 14 | 70% | -1.24 | 8.86 |
|  | >34 | 12 | 7 | 58% | -3.89 | 4.78 | 10 | 6 | 60% | -4.33 | 4.86 |
| **PBS** | < 25 | 21 | 19 | 90% | 4.73 | 3.78 | 16 | 14 | 88% | 6.02 | 4.28 |
|  | 25-34 | 32 | 30 | 94% | 2.61 | 3.99 | 37 | 33 | 89% | 2.01 | 5.08 |
|  | >34 | 9 | 5 | 56% | -5.68 | 3.36 | 14 | 10 | 71% | -7.43 | 4.23 |
| **SSR\*** | < 25 | 14 | 14 | 100% | 3.76 | 4.49 | 7 | 6 | 86% | 3.38 | 3.64 |
|  | 25-34 | 17 | 17 | 100% | 1.18 | 3.38 | 24 | 23 | 96% | 2.17 | 3.9 |
|  | >34 | 12 | 6 | 50% | -0.31 | 4.52 | 15 | 13 | 87% | 1.6 | 3.28 |

\* Sansure sample storage reagent

## Discussion

The standard molecular diagnostic test for SARS-CoV-2 is a multistep process that requires viral RNA extraction and qRT-PCR implementation. Due to increased demand in the rapid and reliable screening of this virus, alternative protocols with similar sensitivity are needed. Efforts to simplify the current methods are critical to assess the viral spread and limit the pandemic, as well as could benefit patients’ care. Recent attempts have been made to omit viral RNA extraction to simplify the direct qRT-PCR protocol of COVID-19 detection and also assess the impact of different swab storage media composition on PCR efficiency [7]–[10]. However, most of the studies are done with small sample size, and needs to be expanded for better understanding performance and caveats associated with direct sample testing,

In this study, we compared different direct qRT-PCR methods (without nucleic acid extraction). Overall, we observed consistent results for SARS-CoV-2 detection, however, the average Ct values for both direct methods were higher than for extracted RNA samples. Our study suggests that heat-inactivation and pelleting have more sensitivity for SARS-CoV-2 detection PCR assay compared to heat-inactivation only. For the HC method +4.65 (for ORF1ab gene) and +5.53 (for N gene) median Ct differences were detected compared to purified nucleic acid samples. This could be for several reasons: i) for the same input volume of samples nucleic acid extraction yields higher quantity/quality of RNA compared with the direct sample; ii) RNA extraction was performed from fresh swab samples, while the aliquots used for the direct method had been stored at +4 °C for at least one day before heat-inactivation and PCR. The storage duration is known to cause the shift of results by 2-3 Ct compared to Ct values for eluates of matched fresh aliquots of the same nasopharyngeal specimens [11].

The overall sensitivity of 88% in our study was comparable to 87.8% (n = 41) reported by Alcoba-Florez *et al*. and was a bit lower than 93% (n = 77) reported by Brown *et al*., however the sample size in both studies were much smaller [5], [9]. We also demonstrated that detection accuracy with direct sample testing highly depends on the concentration of viral particles in the sample, and the reliability of direct testing dramatically decreases in the samples with viral particle concentrations close to the limits of detection.

In conclusion, our results indicate that the detection of SARS-CoV-2 in isolated and purified RNA samples provides more reliable results, and direct methods are characterized by an increase of false-negative detections. On the other side, omitting the isolation step may help to significantly increase testing capacity, and switching to direct sample testing is justified if the number of tests is doubled at least.

## Data Availability

N/A

## Acknowledgments

We are grateful to the National Center for Disease Control of MH RA, National Center for AIDS Prevention of MH RA, and Davidyants Laboratories involved in COVID-19 diagnostic testing for providing nasopharyngeal swabs and RNA samples. We would like to specially thank “Seeding Labs” for supporting this research in the frame of the “Instrumental Access 2017” program.

